# Bridging the Sequencing Gap: N501Y SNP RT-qPCR Assay Detects First SARS-CoV-2 Beta Variant in the Philippines

**DOI:** 10.1101/2024.06.26.24309150

**Authors:** Samantha Louise P. Bado, Niquitta B. Galap, Joanna Ina G. Manalo, Othoniel Jan T. Onza, Mary Rose B. Pelingon, Joy Mariette L. Parayray, Justine Mae Palciso, Karen Iana C. Tomas, Miguel Francisco B. Abulencia, Francisco Gerardo M. Polotan, Lei Lanna M. Dancel, Amalea Dulcene Nicolasora

## Abstract

Whole genome sequencing (WGS) is used extensively in identifying SARS-CoV-2 variants. However, this method requires stringent sample acceptance criteria, long turn-around time (TAT), expensive processing and maintenance costs, and highly skilled staff. Although sequencing offers comprehensive pathogen insights, a cost-effective tool with faster TAT is beneficial in detecting SARS-CoV-2 variants of concern (VOCs). Here, we used a single nucleotide polymorphism (SNP) RT-qPCR assay to detect the N501Y mutation in banked SARS-CoV-2 RNA extracts (N=452) collected from December 2020 to April 2021. Of the SARS-CoV-2 positives (n=367), 22% carried the N501Y mutation and were classified as probable VOCs. This includes a sample that was confirmed to belong to the Beta lineage and was collected earlier than the first reported Beta variant in the country suggesting an earlier emergence of the variant. Validation experiments for the SNP RT-qPCR assay showed a limit of detection (LOD) of 3.01 copies/μL for both N501 and 501Y targets. A 99.35% concordance with partial S gene Sanger sequencing was observed confirming the presence of the N501Y SNP in 83 samples. In conclusion, the optimized SNP RT-qPCR assay serves as an important complementary or alternative tool for detecting probable SARS-CoV-2 variants, ensuring that samples ineligible for WGS are not overlooked. This effectively resolves sequencing gaps, including stringent sample acceptance criteria, extended TAT, and rigorous data analysis. Therefore, embracing this technology provides a rapid, economical, and dependable solution for managing pathogens of public health concern.

## INTRODUCTION

The global emergence of SARS-CoV-2 variants of concern (VOCs) heightened the need for genomic surveillance systems to maintain effective public health interventions during the COVID-19 pandemic(1). In late 2020, an emergence of mutations enhancing viral pathogenicity, transmissibility, and antigenicity were observed(2). Three independent SARS-CoV-2 lineages, B.1.1.7/Alpha(3), B.1.351/Beta(4), and P.1/Gamma(5), were introduced and rapidly multiplied in the population. While these variants are characterized by a myriad of mutations across their genomes, they share the key spike gene mutation N501Y(6). The mutation was identified as a possible determinant of enhanced infection, providing the virus with a more efficient viral entry and the ability to evade anti-spike protein antibodies(6–10).

The World Health Organization (WHO) recommended whole genome sequencing (WGS) for detecting and identifying emerging SARS-CoV-2 variants(11). However, using such technology proves to be challenging in resource-limited countries such as the Philippines due to its expensive processing and maintenance costs. Additionally, the long turnaround time, stringent sample acceptability criteria, and high technical requirements make it disadvantageous, especially in outbreak scenarios. Thus, alternative methods with rapid and efficient workflows are needed to allow SARS-CoV-2 variant screening(12).

An RT-qPCR assay that targets variant-specific SNP mutations like N501Y can be used as a screening tool for early variant classification(11). Laboratory-developed SNP RT-qPCR assays were implemented in the surveillance measures in several countries, which allowed them to screen and identify probable variants rapidly(13–17). In this paper, we optimized and used a SNP RT-qPCR assay to screen for the variant-associated N501Y amino acid substitution in banked SARS-CoV-2 samples. We also describe how it helped identify a missed Beta variant case in the Philippines.

## MATERIALS AND METHODS

### Ethical statement

All samples, protocols, and procedures described in this study are approved by the Research Institute for Tropical Medicine Institutional Review Board (RITM-IRB) with IRB Number 2022-43. The written informed consent requirement was waived based on the context of the emerging outbreak response.

### Clinical specimens

RNA extracts from December 2020 to April 2021 collected from the National Capital Region (NCR) and Region IV-A in the Philippines were acquired from the RITM Advanced Molecular Technologies Laboratory (RITM-AMTL) COVID-19 specimen bank. Four hundred fifty-two (452) archived RNA extracts of PCR-confirmed SARS-CoV-2 negative (n = 85) and positive (n = 367) samples were retrieved and tested at the RITM-AMTL. All patient samples were de-identified and anonymized in all materials and documents used in this study.

### cDNA synthesis

Complementary DNA (cDNA) was synthesized from RNA extracts using Moloney Murine Leukemia Virus (M-MLV) Reverse Transcriptase with RNaseOUT™ Recombinant Ribonuclease Inhibitor and Random Primers according to the manufacturer’s protocol (Invitrogen, CA, USA).

### N501Y SNP RT-qPCR assay

The synthesized cDNA was subsequently tested using the N501Y SNP RT-qPCR assay that targets the N501Y wild type and mutant alleles. All primers and probes (Table 1) were adapted from the National Institute of Infectious Diseases (NIID) of Japan(18). The SNP RT-qPCR mastermix has a total volume of 10μL containing 5μL 2X Taqman Fast Advanced Master Mix (Applied Biosystems, CA, USA), 3.2μL UltraPure™ Distilled Water (Invitrogen, CA, USA), 0.6μM primers, 0.1μM probes, and 1μL cDNA template. The amplification of the N501 and 501Y targets was detected through the FAM and VIC channels, respectively. The thermocycling profile optimized was 60°C for 1 minute, 50°C for 2 minutes, 95°C for 2 minutes, and 40 cycles of 95°C for 3 seconds and 60°C for 30 seconds, then a cycle of 60°C for 1 minute, using the 7500 Fast Real-Time PCR System (Applied Biosystems, CA, USA). Laboratory-confirmed SARS-CoV-2 negatives served as the negative control, and *in vitro* transcribed N501 and 501Y oligonucleotides served as positive controls for each RT-qPCR run.

**Table 1.**
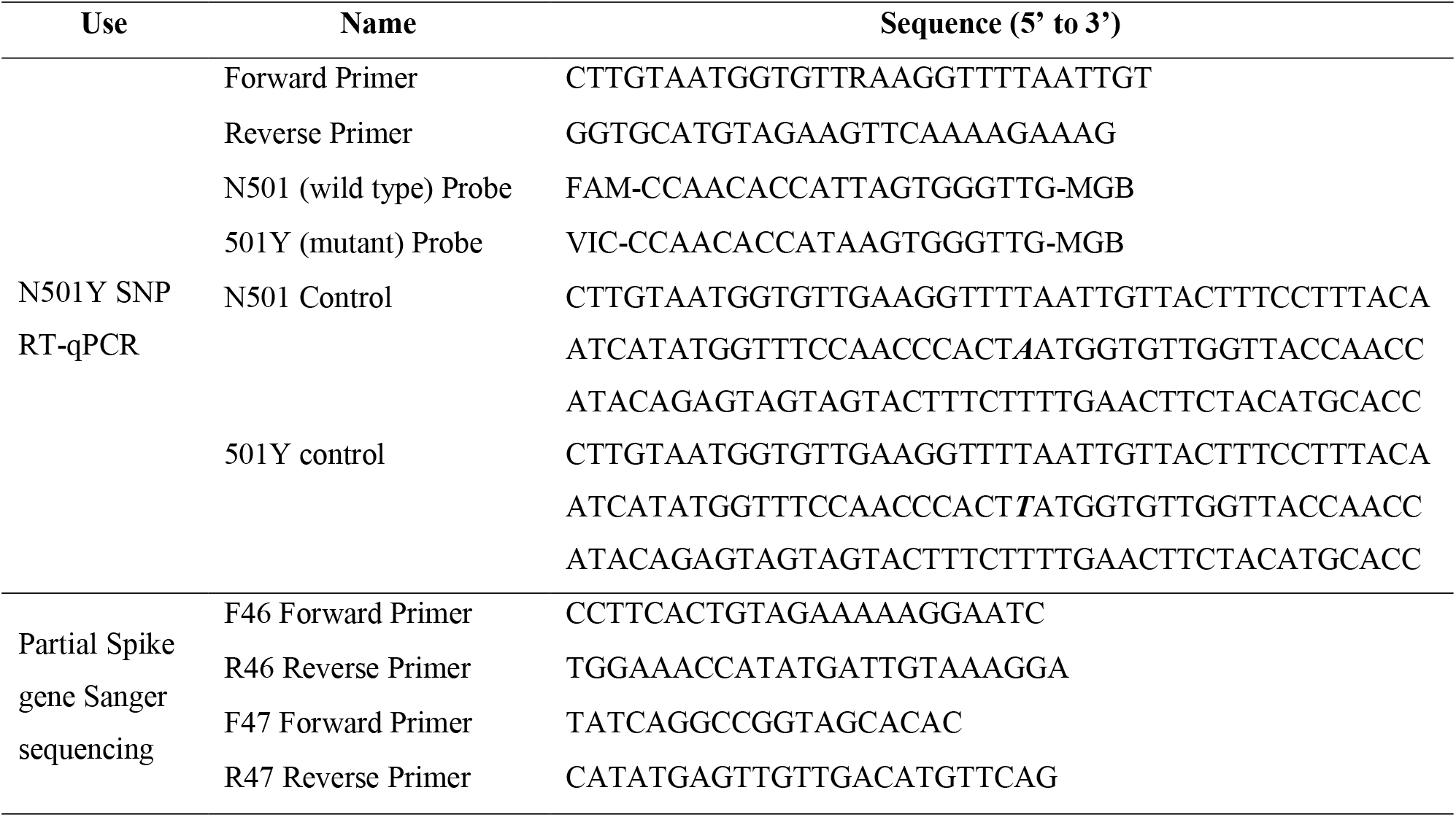
List of primers, probes, and positive control sequences used for N501Y mutation detection.

### N501Y SNP Assay Validation

The SNP assay’s limit of detection (LOD) was validated for both N501 and 501Y targets using *in vitro* transcribed N501 and 501Y oligonucleotides. Starting with 3.01×10^9^ copies/μL, 10-fold serial dilutions were performed down to 3.01×10^−2^ copies/μL. All dilutions were tested in triplicates. The last two dilutions with a 100% detection rate for each target were tested in 20 replicates. The lowest dilution with at least a 95% detection rate is considered the LOD.

A panel of four hundred fifty-two (452) samples that were previously tested for the presence of SARS-CoV-2 using a commercial kit (Sansure Biotech) were used in this study. The SARS-CoV-2-positives were classified according to the Ct values of their ORF1ab gene target during the initial screening test: Low Ct is less than 25, Mid Ct is between 25 and 30, and High Ct is greater than 30. The samples were tested using the N501Y SNP RT-PCR assay to confirm the assay’s viability in detecting SARS-CoV-2 and distinguishing between N501Y wild type and mutant relative to Ct values.

### PCR amplification and Partial Spike Gene Sanger Sequencing

To evaluate the accuracy of the SNP RT-qPCR assay in detecting the N501Y SNP, a subset of 308 SARS-CoV-2-positive samples were sequenced using the Sanger sequencing method. This method is the gold standard for identifying SNPs and basecalling accuracy. Primers adapted from the National Reference Center for Emerging Viral Infections(19) flanking the Spike gene region were used to amplify two overlapping fragments covering positions 914-1981 of the Spike gene.

The conventional PCR reactions were carried out using the Ex Taq® PCR kit (TaKaRa, Kusatsu, Japan). The first round PCR mastermix has a total volume of 20μL containing the F46 and R47 primers at 500nM each, 2.0μL of 10x Ex Taq buffer, 1.6μL of 10μM dNTP mix, 0.1μL of Taq enzyme, and 0.1μL cDNA. Pre-denaturation was set at 94°C for 2 minutes, followed by 35 cycles of denaturation at 94°C for 30 seconds, annealing at 55°C for 30 seconds, and extension at 72°C for 1 minute, then followed by a final extension at 72°C for 10 minutes. Two separate hemi-nested PCR runs were performed, one using F46 and R46 primers and the other using F47 and R47 primers (see Table 1). Each reaction mix consists of 500nM each of the primers, 2.0μL of 10x Ex Taq buffer, 1.6μL of 10μM dNTP mix, 0.1μL of Taq enzyme, and 0.5 μL of template with a total volume of 20μL. The PCR profile from the first round was altered for the nested round by reducing the extension stage from 1 minute to 30 seconds at 72°C. The PCR products were visualized using agarose gel electrophoresis. Samples with clear, distinct bands were purified using PureLink PCR purification kit (Invitrogen, CA, USA) following the manufacturer’s protocol. Samples were sent to a third-party sequencing facility for sequencing using the ABI 3730XL system.

### Sequence analysis

Partial S gene sequences were trimmed and assembled using MEGA X and Aliview software(20,21), and alignment was done using the S gene reference sequence (EPI_ISL_402124). Consensus FASTA sequences were uploaded to the Stanford Coronavirus Antiviral & Resistance Database (CoVDB) to check for the presence of the N501Y mutation. The program uses the reference sequence Wuhan-Hu-1 (NC_045512.2) for alignment(22).

### Phylogenetic Analysis

The N501Y-containing sample with the earliest collection date, PH-RITM-0142, was sequenced through a modified ARTIC protocol(23). Fifty-seven (57) of its closely related sequences from the Philippines (EPI_SET_230914kr) were obtained using Audacity*Instant* (V5.1.0) on GISAID(24) on September 12, 2023. The sequences were aligned using MAFFT v7.520(25) and a phylogenetic tree was generated using IQ-TREE v2.2.3(26) with a GTR nucleotide substitution model and a fixed clock rate. The resulting tree was input into TreeTime v0.10.1(27) with 1000 iterations for bootstrapping to generate a time-scaled phylogeny.

## RESULTS

### N501Y SNP RT-qPCR Assay Validation

The SNP RT-qPCR assay’s limit of detection (LOD) was validated to be 3.01 copies/μL for both N501 and 501Y targets with 100% and 95% detection rates in 20 replicates, respectively (see Table S1). All SARS-CoV-2 negative samples (n=85) also showed negative results using the SNP assay. Among the SARS-CoV-2-positives, 48 (13%) showed negative results and were not classified by the SNP RT-qPCR assay as either wild type or mutant as shown in Figure 1. Of these 48 samples, one had a mid-range Ct value and 47 exhibited a high Ct value for the SARS-CoV-2 PCR screening test. Figure 1 illustrates a decreasing trend in the detection rate of the SNP RT-qPCR assay as the Ct value of the samples increases. The concordance rate between the commercial kit (Sansure Biotech) and the SNP RT-qPCR assay is 100% and 87% in SARS-CoV-2 negative and positive samples.

**Figure 1.**
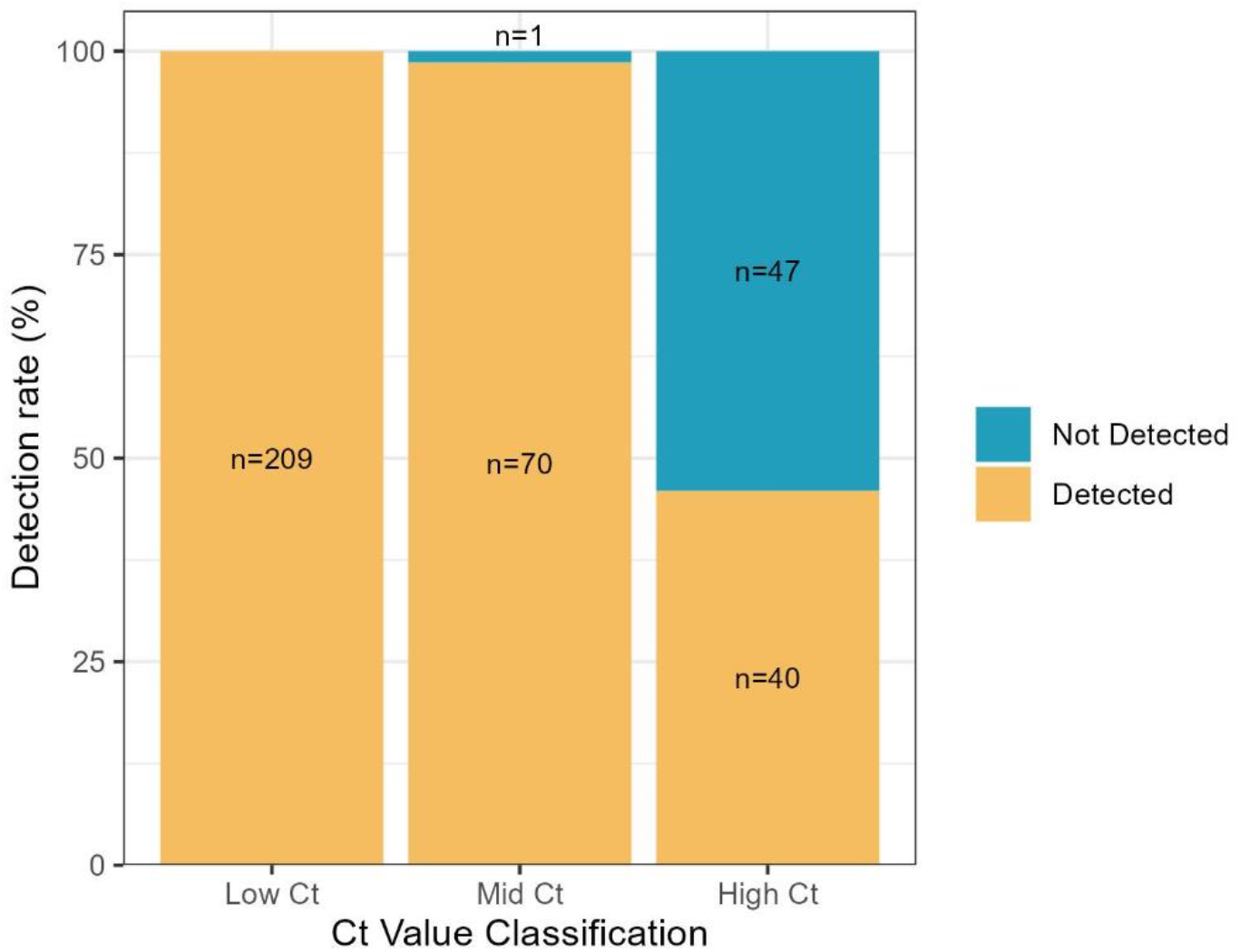
SARS-CoV-2 detection rate of N501Y SNP RT-qPCR assay based on Ct value generated from the ORF1ab gene target of the Sansure RT-qPCR run. The samples were classified based on the following scheme: Low Ct <25, Mid Ct 25-30, and High Ct >30.

Concordance testing with partial S gene Sanger sequencing using 308 SARS-CoV-2-positive samples (225 wild types and 83 with N501Y mutation) shows an overall percent agreement (OPA) of 99.35% where the SNP RT-qPCR assay correctly classified 306 samples. Only two samples containing the 501Y mutation according to Sanger sequencing exhibited discordant results (see Table 2).

**Table 2.** Concordance testing results between N501Y SNP RT-qPCR assay and partial S gene Sanger sequencing.

| SNP detection | Sanger mutant (501Y) | Sanger wild type (N501) | Total |
| --- | --- | --- | --- |
| SNP detected (501Y) | 81 | 0 | 81 |
| SNP not detected (N501) | 2 | 225 | 227 |
| Total | 83 | 225 | 308 |

### Comparing the SARS-CoV-2 lineage frequency distribution using SNP RT-qPCR vs Whole Genome Sequencing

Of the 452 samples subjected to N501Y SNP RT-qPCR, a shift in the proportion of samples without the N501Y mutation (N501) to those with the N501Y mutation (501Y) was observed between December 2020 and April 2021 (Figure 2A). The detection rate of N501Y mutants increased by 95.32% from December 2020 to April 2021. The increasing trend coincides with that of Philippine whole genome sequences as shown in Figure 2B. However, it can be noted that the detection of the spike mutation is earlier for the SNP RT-qPCR (December 2020) as compared to WGS (January 2021). Notably, one of the samples that carried the N501Y mutation, PH-RITM-0142, was later confirmed by WGS to be a Beta variant. The sample was collected on December 9, 2020, which is earlier than the first reported Beta case in the country.

**Figure 2.**
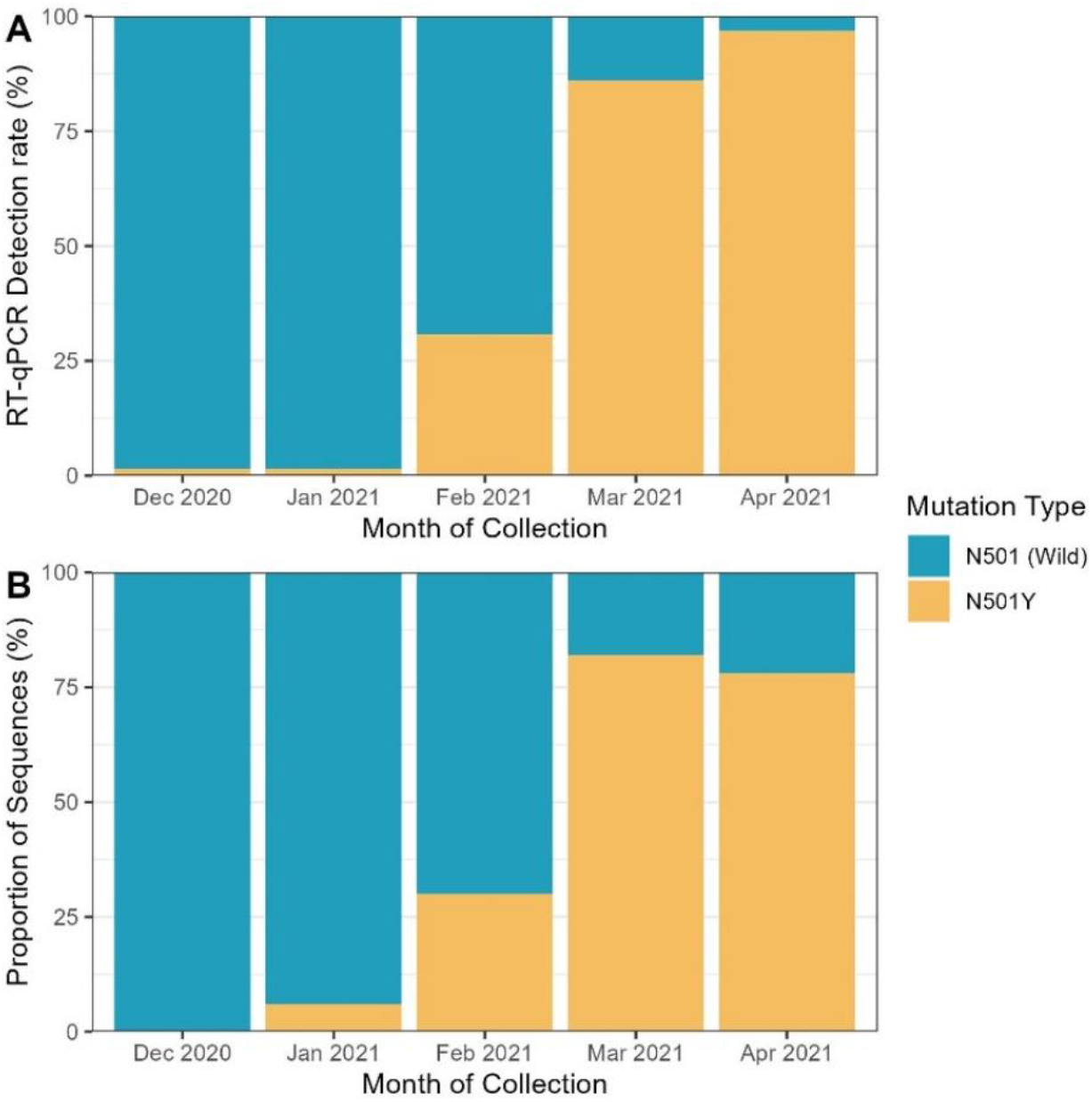
(A) Detection rate of N501 and 501Y among SARS-CoV-2 positive samples from December 2020 to April 2021 using SNP RT-qPCR, (B) Philippine whole genome sequences collected between December 2020 and April 2021 carrying the spike mutation.

The prevalence of the N501Y mutation among tested samples aligns with the increase in confirmed cases of SARS-CoV-2 and the shift from the predominantly circulating lineage B.1.1.63 to the Alpha and Beta variants (Figure 3). However, sequencing data suggests that the first Beta variant cases were only introduced in the country around February 2021, which contrasts with our identification of a Beta variant sampled in December 2020.

**Figure 3.**
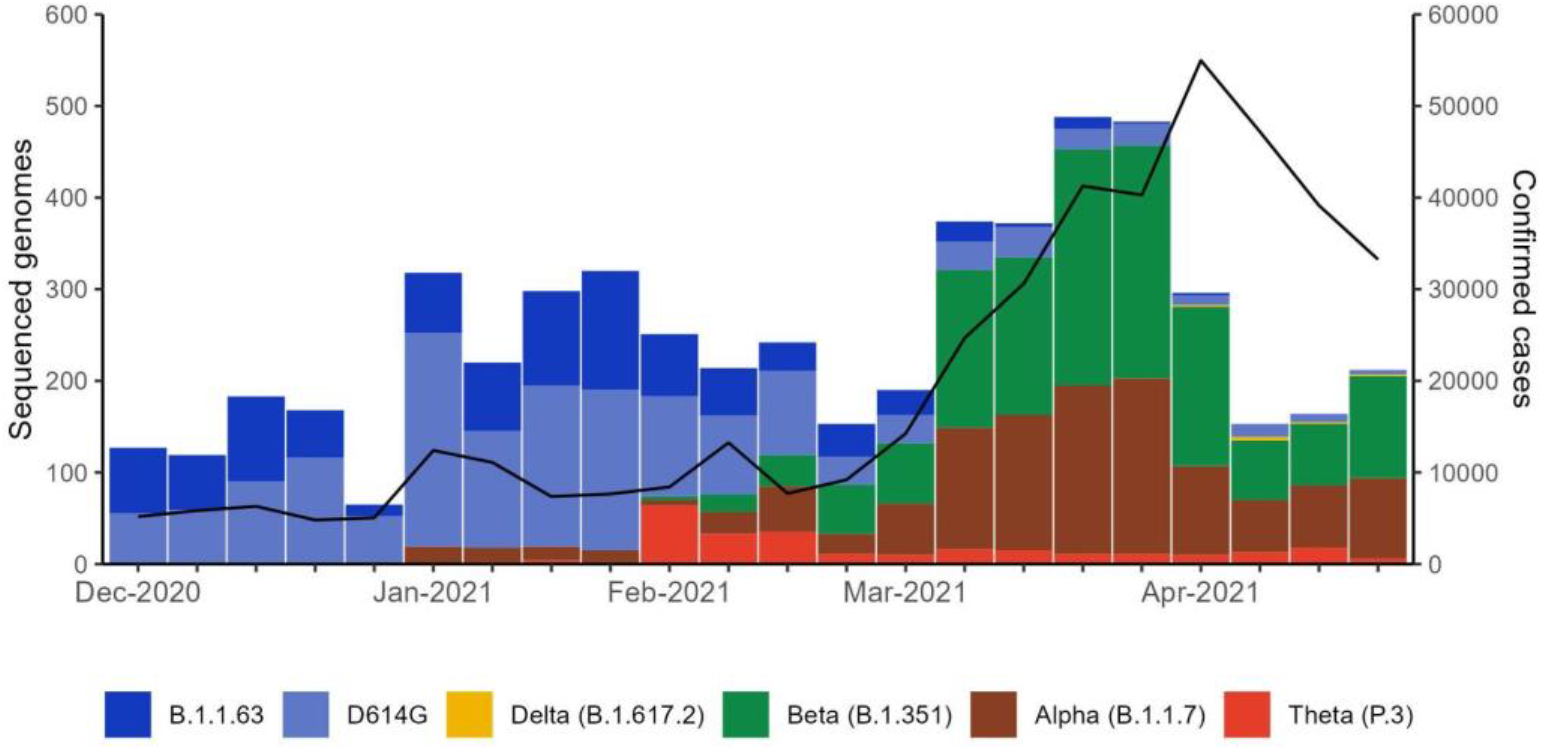
Number of sequenced genomes for each major variant (bars) and confirmed COVID-19 cases (black solid line). D614G corresponds to non-major variants carrying the D614G mutation.

### Phylogenetic Analysis

Fifty-seven (57) B.1.351/Beta sequences collected in the Philippines that were closely related to PH-RITM-0142 were generated using Audacity*Instant* (V5.1.0) on GISAID. In Figure 4, phylogenetic analysis revealed that the PH-RITM-0142 B.1.351/Beta variant was introduced earlier than what was reported to be the first case.

**Figure 4.**
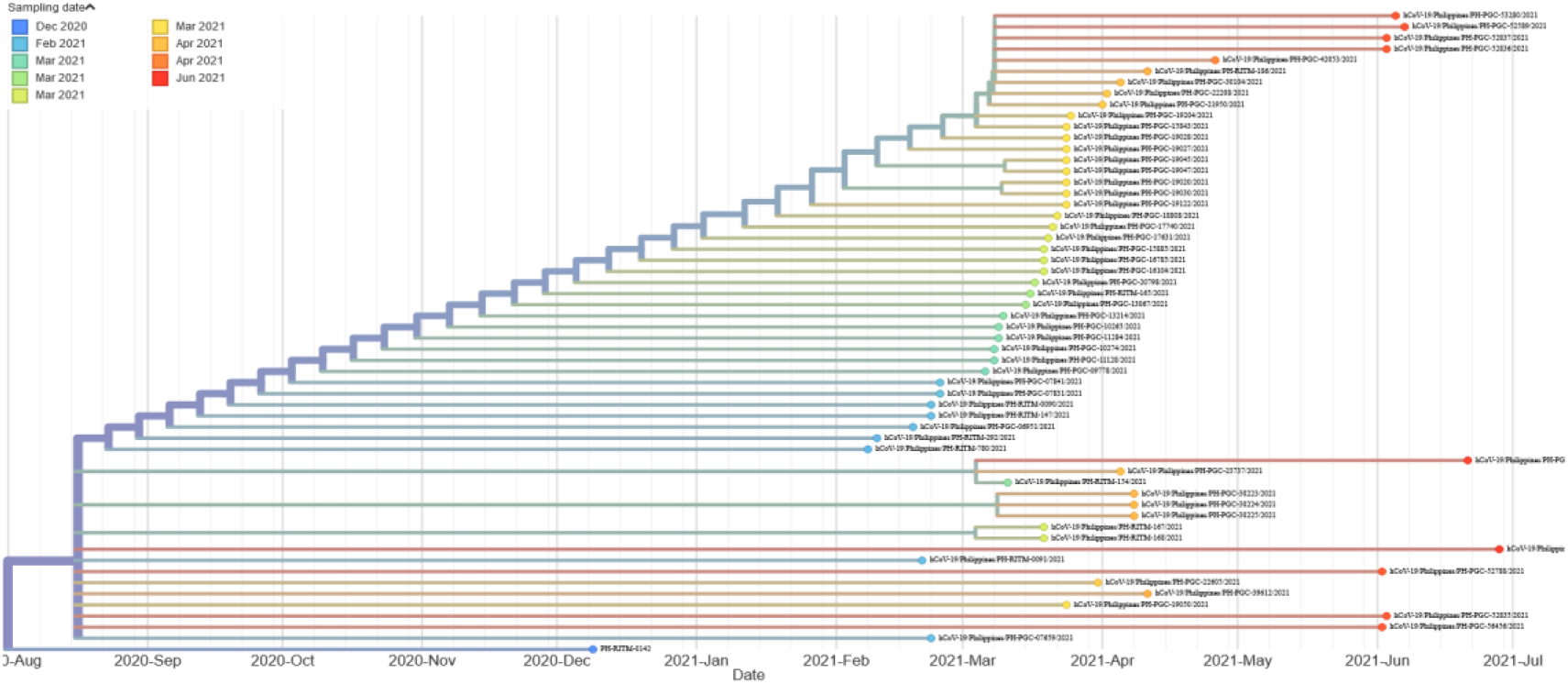
Phylogenetic analysis of the first N501Y case - PH-RITM-0142 - and its closest sequences collected in the Philippines.

## DISCUSSION

This study documents how we developed, optimized, and validated a SNP RT-qPCR assay to detect the N501Y mutation in archived SARS-CoV-2 samples collected from NCR and Region IV-A in the Philippines from December 2020 to April 2021. Our findings demonstrate its potential use in public health emergency response. N501Y is a key spike gene mutation shared by the Alpha, Beta, and Gamma variants(28), which were VOCs circulating globally at the time of collection of the archived samples. The first reported VOC case in the Philippines belonged to the Alpha lineage and was reported on January 13, 2021(29,30), although current data from GISAID shows that the Alpha variant was present as early as December 10, 2020 (EPI_ISL_1081795; https://doi.org/10.55876/gis8.240515yr). Data also shows that the first Beta case detected in the country was on January 31, 2021 (EPI_ISL_4743768; https://doi.org/10.55876/gis8.240515ay). Interestingly, in testing the archived samples, we detected the N501Y mutation from a sample collected on December 9, 2020, later confirmed by WGS to be of B.1.351/Beta lineage. The sample initially did not meet the stringent criteria for WGS since its ORF1ab Ct value is 32.63, which is beyond the recommended Ct value of ≤30(31). This case highlights a gap in variant surveillance efforts, where samples with Ct values exceeding the eligibility criteria for WGS might be excluded from sequencing, potentially leading to the missed detection of important variants. In acknowledgment of such instances, non-sequencing methods like our optimized PCR-based SNP (N501Y) detection assay become a crucial source of information for rapid variant detection.

Our optimized SNP RT-qPCR assay demonstrates high concordance with partial S gene Sanger sequencing in detecting the N501Y SNP mutation. These findings signify the potential impact of utilizing rapid detection methods, in complementary with sequencing methodologies, in the quick estimation of the prevalence of probable variants in the community to help facilitate immediate preventive measures against its spread.

Analytical performance validation of our SNP RT-qPCR assay showed an LOD of 3.01 copies/μL for both targets in the validation experiments. When compared with a commercial SARS-CoV-2 diagnostic kit that targets the N and ORF1ab SARS-CoV-2 genes in a panel of clinical respiratory samples with low-intermediate- and high-concentration of SARS-CoV-2 genetic material, our in-house SNP (N501Y) assay could only detect SARS-CoV-2 from 46% of samples with Ct values >30 (estimated to have low SARS-CoV-2 genetic material concentration) and subsequently classify them as either wild type or mutant for the N501Y mutation. While this indicates that our SNP assay has a lower analytical sensitivity than the comparator commercial SARS-CoV-2 diagnostic kit whose LOD is 0.2 copies/μL(32), we emphasize that the assay is not meant to be used as a standalone diagnostic test for SARS-CoV-2. Its primary utility is for detecting the target SNP, in this case, the N501Y mutation.

In terms of detecting the N501Y SNP, the assay exhibited high diagnostic specificity and sensitivity with an overall percent agreement (OPA) of 99.35% relative to Sanger sequencing. These results show that the assay is sufficiently specific and sensitive in detecting the N501Y mutation. This makes it a useful tool for screening for probable variants, followed by sequencing as a confirmatory step.

Our retrospective testing revealed that B.1.351/Beta was detected in a sample (PH-RITM-0142) collected on December 9, 2020, from a high-risk patient with no known exposure and travel history. This finding suggests earlier circulation of the Beta variant compared to the officially reported date of March 2, 2021, which was based on the earliest confirmed case at that time, whose sample was collected in late January(33). This also shows that SARS-CoV-2 VOCs were already circulating in the country before its first public report in January 2021. Initially, the sample was not included in the subset that was routinely sequenced for surveillance since its RT-qPCR results showed a high Ct value (32.63) indicating weak positivity. However, since the SNP RT-qPCR results showed that it carried the N501Y mutation, we subjected it to WGS using Oxford Nanopore Technology (ONT) for confirmation. Results showed that the sample belonged to the B.1.351/Beta lineage with 81% genome coverage. Moreover, our phylogenetic analysis in Figure 4 further supports the claim that PH-RITM-0142 predates the Beta transmission window in the Philippines, which occurred from January to August 2021 with the first officially reported first case on January 31, 2021. Additionally, it is not linked to the subsequent increase in B.1.351/Beta cases. These results are crucial for determining when and how the B.1.351/Beta variant entered and spread in the Philippines. It suggests that there was an introduction of Beta earlier than initially thought.

While genomic sequencing is important for surveillance, it is still limited to specialized and highly equipped laboratories. It also requires high quality and concentration of SARS-CoV-2 viral RNA as a quality control checkpoint. In the context of outbreak response, a PCR-based method that can screen for probable variants and be deployed more easily might have a greater impact on disease control measures. A multi-step screening concept was developed across diagnostic and research laboratories in Switzerland to understand the spread of VOC and adapt public health interventions. This includes microbiological risk definition using S-dropout as an indicator for the Alpha lineage, and the establishment of N501Y-specific RT-PCR assays(15). Another research initiative opted to use an RT-qPCR screening test, facilitating the analysis of a larger sample size within a shorter timeframe and at a reduced cost(16). They identified variants in samples positive for SARS-CoV-2 by an RT-qPCR mutation screening kit and subsequently confirmed through Next Generation Sequencing (NGS). During the early stages of the COVID-19 pandemic in Argentina, variant surveillance was initiated using WGS. In February 2021, they changed their strategy to partial S gene sequencing due to a significant increase in demand for WGS. However, both sequencing approaches were time-consuming which resulted in delayed identification of variants. They then established an algorithm using an RT-qPCR strategy that targeted the N501Y, E484K, and L452 mutations(17). Similarly, our optimized assay would have been a useful tool for more rapid identification of variants in the country had it been included in our surveillance initiatives as a screening step. Our findings provide use-case evidence of how the limitations of WGS have a big impact on generating an accurate picture of lineage distribution and replacement events. Additionally, it provides evidence of the utility of SNP RT-qPCR assays for variant detection and surveillance in tandem with WGS.

Despite its remarkable diagnostic performance, our optimized assay does not differentiate between the VOCs that share the N501Y SNP which warrants a confirmatory sequencing step to determine the sample’s lineage. Moreover, the assay only targets the N501Y SNP which makes it inapplicable for emerging SARS-CoV-2 VOCs that do not carry the SNP. Additional targets may be included similar to other assays that have already been developed(34–36) to screen for other circulating variants. There is a need to continually update the SNP RT-qPCR assays to accommodate EREIDs that rapidly evolve. We only tested archived samples from NCR and Region IV with the earliest collection date of December 2020. While the assay was able to uncover a shift in the N501Y SNP’s detection rate, we may not have captured potential earlier cases predating December 2020 from other regions. This limits our comprehension of the introductory events of VOCs in the country.

Using rapid tools for detecting VOCs is critical, especially during their early stage of spread. As demonstrated by the detection of an unreported earlier VOC case, PCR-based assays offer the fastest and most practical approach for controlling the spread of new variants which warrants prioritization of detection over depth of information. Although currently limited to N501Y mutations, the optimized SNP RT-qPCR assay can be customized to detect other critical mutations, preparing us to identify emerging viral strains with higher public health significance. Integrating these assays into testing algorithms for outbreak response offers a valuable tool for the immediate implementation of efficient evidenced-based control measures. This proactive approach strengthens our preparedness and response to emerging infectious diseases.

## CONCLUSIONS

In testing archived samples using the optimized highly sensitive and specific SNP RT-qPCR assay, we detected an earlier Beta variant case in the country that was previously missed because the sample failed to meet the stringent WGS eligibility criteria. As an alternative to the labor- and resource-intensive WGS method, or as a complementary method for variant detection, the assay offers a cheap and rapid way to screen for probable variants and estimate their prevalence in the community. We demonstrate here that PCR-based SNP variant detection assays can be an effective complementary tool in assembling a more accurate picture of SARS-CoV-2 transmission. This highlights its utility for rapid outbreak response and the conduct of subsequent surveillance for emerging SARS-CoV-2 variants. Moreover, its adaptability is valuable in addressing potential outbreaks caused by novel pathogens. Our study also highlights the importance of developing and maintaining in-house capacity for the development of molecular assays that can promptly be adjusted to suit the evolving testing needs given the volatile nature of emerging and re-emerging infectious diseases. As we navigate the evolving challenges of infectious diseases, the importance of the adaptability and effectiveness of these assays becomes apparent, ensuring we remain at the lead in the continuous fight against emerging pathogens.

## ACKNOWLEDGMENT

We sincerely thank all the data contributors, namely the authors and their originating laboratories responsible for obtaining the specimens, as well as the submitting laboratories for generating the genetic sequences and metadata. This research is based on their valuable contributions through the GISAID Initiative. We would also like to acknowledge Aldrin V. Imbag and Fatima Rose S. Guemo for maintaining the HPC cluster used in this study. The preparation of this manuscript benefited from guidance and discussion at Project BUKLOD’s Academic Writing Course in June 2023 (funded by the German Alliance for Global Health Research).

## SUPPLEMENTAL DATA

**Table S1.** Limit of detection raw data.

| Replicate | N501 |  | 501Y |  |
| --- | --- | --- | --- | --- |
| | 3.01 copies/μL | 0.301 copies/μL | 3.01 copies/μL | 0.301 copies/μL |
| 1 | Detected | Not Detected | Detected | Detected |
| 2 | Detected | Not Detected | Detected | Not Detected |
| 3 | Detected | Not Detected | Detected | Detected |
| 4 | Detected | Not Detected | Detected | Not Detected |
| 5 | Detected | Not Detected | Detected | Detected |
| 6 | Detected | Detected | Detected | Detected |
| 7 | Detected | Not Detected | Detected | Detected |
| 8 | Detected | Not Detected | Detected | Detected |
| 9 | Detected | Not Detected | Detected | Detected |
| 10 | Detected | Not Detected | Detected | Detected |
| 11 | Detected | Not Detected | Detected | Not Detected |
| 12 | Detected | Detected | Detected | Not Detected |
| 13 | Detected | Not Detected | Detected | Detected |
| 14 | Detected | Not Detected | Detected | Not Detected |
| 15 | Detected | Not Detected | Not Detected | Not Detected |
| 16 | Detected | Detected | Detected | Not Detected |
| 17 | Detected | Detected | Detected | Not Detected |
| 18 | Detected | Not Detected | Detected | Not Detected |
| 19 | Detected | Not Detected | Detected | Detected |
| 20 | Detected | Not Detected | Detected | Detected |
| No. of Replicates | 20 | 20 | 20 | 20 |
| No. of positives | 20 | 4 | 19 | 11 |
| **% Detected** | **100** | **20** | **95** | **55** |

### Data Availability

GISAID Identifier: EPI_SET_230914kr

https://doi.org/10.55876/gis8.230914kr

All genome sequences and associated metadata in this dataset are published in GISAID’s EpiCoV database. To view the contributors of each individual sequence with details such as accession number, Virus name, Collection date, Originating Lab and Submitting Lab, and the list of Authors, visit 10.55876/gis8.230914kr

### Data Snapshot

- EPI_SET_230914kr is composed of 57 individual genome sequences.
- The collection dates range from 2021-02-08 to 2021-06-28;
- Data were collected in 1 countries and territories;
- All sequences in this dataset are compared relative to hCoV-19/Wuhan/WIV04/2019 (WIV04), the official reference sequence employed by GISAID (EPI_ISL_402124). Learn more at https://gisaid.org/WIV04.

## REFERENCES

1. WHO. Global genomic surveillance strategy for pathogens with pandemic and epidemic potential, 2022–2032. 2022; Available from: https://www.who.int/publications-detail-redirect/9789240046979

2. Harvey WT, Carabelli AM, Jackson B, Gupta RK, Thomson EC, Harrison EM, et al. SARS-CoV-2 variants, spike mutations and immune escape. Nat Rev Microbiol. 2021 Jul;19(7):409–24.

3. Rambaut A, Loman N, Pybus O, Barclay W, Barrett J, Carabelli A, et al. Preliminary genomic characterisation of an emergent SARS-CoV-2 lineage in the UK defined by a novel set of spike mutations [Internet]. 2020 Dec. Available from: https://virological.org/t/preliminary-genomic-characterisation-of-anemergent-sars-cov-2-lineage-in-the-uk-defined-by-a-novel-set-of-spike-mutations/563

4. Tegally H, Wilkinson E, Giovanetti M, Iranzadeh A, Fonseca V, Giandhari J, et al. Detection of a SARS-CoV-2 variant of concern in South Africa. Nature. 2021 Apr 15;592(7854):438–43.

5. Faria NR, Mellan TA, Whittaker C, Claro IM, Candido DDS, Mishra S, et al. Genomics and epidemiology of the P.1 SARS-CoV-2 lineage in Manaus, Brazil. Science. 2021 May 21;372(6544):815–21.

6. Leung K, Shum MH, Leung GM, Lam TT, Wu JT. Early transmissibility assessment of the N501Y mutant strains of SARS-CoV-2 in the United Kingdom, October to November 2020. Eurosurveillance [Internet]. 2021 Jan 7 [cited 2023 Sep 27];26(1). Available from: https://www.eurosurveillance.org/content/10.2807/1560-7917.ES.2020.26.1.2002106

7. Tian F, Tong B, Sun L, Shi S, Zheng B, Wang Z, et al. N501Y mutation of spike protein in SARS-CoV-2 strengthens its binding to receptor ACE2. eLife. 2021 Aug 20;10:e69091.

8. Williams AH, Zhan CG. Fast Prediction of Binding Affinities of the SARS-CoV-2 Spike Protein Mutant N501Y (UK Variant) with ACE2 and Miniprotein Drug Candidates. J Phys Chem B. 2021 May 6;125(17):4330–6.

9. Liu Y, Liu J, Plante KS, Plante JA, Xie X, Zhang X, et al. The N501Y spike substitution enhances SARS-CoV-2 transmission [Internet]. Microbiology; 2021 Mar [cited 2023 Nov 30]. Available from: http://biorxiv.org/lookup/doi/10.1101/2021.03.08.434499

10. Sandoval Torrientes M, Castelló Abietar C, Boga Riveiro J, Álvarez-Argüelles ME, Rojo-Alba S, Abreu Salinas F, et al. A novel single nucleotide polymorphism assay for the detection of N501Y SARS-CoV-2 variants. J Virol Methods. 2021 Aug;294:114143.

11. ECDC, WHO. Methods for the detection and characterisation of SARS-CoV-2 variants - second update [Internet]. 2022 Aug. Available from: https://www.ecdc.europa.eu/sites/default/files/documents/Methods-for-the-detection-char-SARS-CoV-2-variants_2nd%20update_final.pdf

12. Specchiarello E, Matusali G, Carletti F, Gruber CEM, Fabeni L, Minosse C, et al. Detection of SARS-CoV-2 Variants via Different Diagnostics Assays Based on Single-Nucleotide Polymorphism Analysis. Diagnostics. 2023 Apr 27;13(9):1573.

13. Abdulnoor M, Eshaghi A, Perusini SJ, Broukhanski G, Corbeil A, Cronin K, et al. Real-Time RT-PCR Allelic Discrimination Assay for Detection of N501Y Mutation in the Spike Protein of SARS-CoV-2 Associated with B.1.1.7 Variant of Concern. Bard JD, editor. Microbiol Spectr. 2022 Feb 23;10(1):e00681–21.

14. Gomes L, Jeewandara C, Jayadas TP, Dissanayake O, Harvie M, Guruge D, et al. Surveillance of SARS-CoV-2 variants of concern by identification of single nucleotide polymorphisms in the spike protein by a multiplex real-time PCR. J Virol Methods. 2022 Feb;300:114374.

15. Goncalves Cabecinhas AR, Roloff T, Stange M, Bertelli C, Huber M, Ramette A, et al. SARS-CoV-2 N501Y Introductions and Transmissions in Switzerland from Beginning of October 2020 to February 2021—Implementation of Swiss-Wide Diagnostic Screening and Whole Genome Sequencing. Microorganisms. 2021 Mar 25;9(4):677.

16. Muñoz-Valle JF, Venancio-Landeros AA, Sánchez-Sánchez R, Reyes-Díaz K, Galindo-Ornelas B, Hérnandez-Monjaraz WS, et al. An Upgrade on the Surveillance System of SARS-CoV-2: Deployment of New Methods for Genetic Inspection. Int J Mol Sci. 2022 Mar 15;23(6):3143.

17. Castro GM, Sicilia P, Bolzon ML, Lopez L, Barbás MG, Pisano MB, et al. Tracking SARS-CoV-2 Variants Using a Rapid Typification Strategy: A Key Tool for Early Detection and Spread Investigation of Omicron in Argentina. Front Med. 2022 May 17;9:851861.

18. Miyamoto S, Arashiro T, Ueno A, Kanno T, Saito S, Katano H, et al. Non-Omicron breakthrough infection with higher viral load and longer vaccination-infection interval improves SARS-CoV-2 BA.4/5 neutralization. iScience. 2023 Feb;26(2):105969.

19. Université de Genève, Hôpitaux Universitaires de Genève. Protocol for specific RT-PCRs for marker regions of the Spike region indicative of the UK SARS-CoV2 variant B.1.1.7 and the South African variant 501Y.V2 [Internet]. 2020 Dec. Available from: https://www.hug.ch/sites/interhug/files/structures/laboratoire_de_virologie/protocol_amplification_voc_20201201_uk_geneva.pdf

20. Larsson A. AliView: a fast and lightweight alignment viewer and editor for large datasets. Bioinformatics. 2014 Nov 15;30(22):3276–8.

21. Kumar S, Stecher G, Li M, Knyaz C, Tamura K. MEGA X: Molecular Evolutionary Genetics Analysis across Computing Platforms. Battistuzzi FU, editor. Mol Biol Evol. 2018 Jun 1;35(6):1547–9.

22. Stanford Coronavirus Antiviral & Resistance Database (CoVDB) [Internet]. [cited 2023 Oct 9]. Available from: https://covdb.stanford.edu/sierra/sars2/by-sequences/

23. Quick J. nCoV-2019 sequencing protocol v1 [Internet]. 2020 [cited 2024 Jun 18]. Available from: https://www.protocols.io/view/ncov-2019-sequencing-protocol-bbmuik6w

24. Shu Y, McCauley J. GISAID: Global initiative on sharing all influenza data – from vision to reality. Eurosurveillance [Internet]. 2017 Mar 30 [cited 2023 Oct 4];22(13). Available from: https://www.eurosurveillance.org/content/10.2807/1560-7917.ES.2017.22.13.30494

25. Katoh K. MAFFT: a novel method for rapid multiple sequence alignment based on fast Fourier transform. Nucleic Acids Res. 2002 Jul 15;30(14):3059–66.

26. Nguyen LT, Schmidt HA, Von Haeseler A, Minh BQ. IQ-TREE: A Fast and Effective Stochastic Algorithm for Estimating Maximum-Likelihood Phylogenies. Mol Biol Evol. 2015 Jan;32(1):268–74.

27. Sagulenko P, Puller V, Neher RA. TreeTime: Maximum-likelihood phylodynamic analysis. Virus Evol [Internet]. 2018 Jan 1 [cited 2023 Nov 30];4(1). Available from: http://academic.oup.com/ve/article/doi/10.1093/vex042/4794731

28. Young M, Crook H, Scott J, Edison P. Covid-19: virology, variants, and vaccines. BMJ Med. 2022 Mar;1(1):e000040.

29. DOH. PH Genomic Biosurveillance Detects SARS-CoV-2 UK Variant [Internet]. 2021 Jan. Available from: https://doh.gov.ph/doh-press-release/PH-GENOMIC-BIOSURVEILLANCE-DETECTS-SARS-COV-2-UK-VARIANT

30. PGC. PGC SARS-CoV-2 Bulletin No. 6: First case of the new variant under Lineage B.1.1.7 detected in the Philippines [Internet]. 2021 Jan. Available from: https://pgc.up.edu.ph/pgc-sars-cov-2-bulletin-no-6/

31. Pan American Health Organization. Guidance for SARS-CoV-2 samples selection for genomic characterization and surveillance [Internet]. 2021. Available from: https://www.paho.org/en/file/82020/download?token=dUPSGHUm

32. Sansure Biotech, Inc. Novel Coronavirus (2019-nCoV) Nucleic Acid Diagnostic Kit (PCR-Fluorescence Probing) Instructions for Use V02. 2022.

33. Ropero G. PH reports 10,019 more COVID-19 cases, total now at 2,632,881. ABS-CBN News. 2021 Oct 7;

34. Martinez M, Nguyen PV, Su M, Cardozo F, Valenzuela A, Franco L, et al. SARS-CoV-2 Variants in Paraguay: Detection and Surveillance with an Economical and Scalable Molecular Protocol. Viruses. 2022 Apr 22;14(5):873.

35. Nörz D, Grunwald M, Tang HT, Olearo F, Günther T, Robitaille A, et al. Rapid Automated Screening for SARS-CoV-2 B.1.617 Lineage Variants (Delta/Kappa) through a Versatile Toolset of qPCR-Based SNP Detection. Diagnostics. 2021 Oct 1;11(10):1818.

36. Wang H, Miller JA, Verghese M, Sibai M, Solis D, Mfuh KO, et al. Multiplex SARS-CoV-2 Genotyping Reverse Transcriptase PCR for Population-Level Variant Screening and Epidemiologic Surveillance. McAdam AJ, editor. J Clin Microbiol. 2021 Jul 19;59(8):e00859–21.

